# Acute kidney injury in hospitalized patients due to COVID-19

**DOI:** 10.1101/2021.05.31.21257656

**Authors:** Pehuén Fernández, Emanuel José Saad, Augusto Douthat, Federico Ariel Marucco, María Celeste Heredia, Ayelén Tarditi Barra, Silvina Trinidad Rodriguez Bonazzi, Melani Zlotogora, María Antonella Correa Barovero, Sofía Villada, Juan Pablo Maldonado, Juan Pablo Caeiro, Ricardo Arturo Albertini, Jorge De La Fuente, Walter Guillermo Douthat

## Abstract

The incidence of acute kidney injury (AKI) in hospitalized patients with coronavirus disease 2019 (COVID-19) is variable, being associated with worse outcomes. The objectives of the study were to evaluate the incidence, risk factors and impact of AKI in subjects hospitalized for COVID-19 in two third- level hospitals in Córdoba, Argentina.

A retrospective cohort study was conducted. 448 adults who were consecutively hospitalized for COVID-19 between March and the end of October 2020 at Hospital Privado Universitario de Córdoba and Hospital Raúl Angel Ferreyra were included. The incidence of AKI was 19% (n = 85). 50.6% presented AKI stage 1 (n=43), 20% stage 2 (n=17) and 29.4% stage 3 (n=25, of which 18 required renal replacement therapy). In the multivariate analysis, the variables that were independently associated with AKI were: age (adjusted Odd ratio -aOR- =1.30, 95%CI=1.04-1.63, p=0.022), history of chronic kidney disease (aOR=9.92, 95%CI=4.52-21.77, p<0.001), blood neutrophil count at admission (aOR=1.09, 95%CI=1.01-1.18, p=0.037) and requirement for mechanical ventilation (MV) (aOR=6.69, 95%CI=2.24-19.9, p=0.001). AKI was associated with longer hospitalization, greater admission and length of stay in the intensive care unit, a positive association with bacterial superinfection, sepsis, respiratory distress syndrome, MV requirement and mortality (mortality with AKI=47.1% vs without AKI=12.4%, p<0.001). AKI was independently associated with higher mortality (aOR=3.32, 95%CI=1.6-6.9, p=0.001).

In conclusion, the incidence of AKI in adults hospitalized for COVID-19 was 19% and had a clear impact on morbidity and mortality. Local predisposing factors for AKI were identified.

## Introduction

The outbreak of severe acute respiratory syndrome (SARS-CoV-2) due to coronavirus 2019 (COVID-19) has quickly turned into a global pandemic with devastating consequences^1,2^. The first case reported in Córdoba, Argentina was on March 3, 2020^3^. As of May 8, 2021, in Córdoba more than 230 thousand infected people had been reported, and more than 3.1 million in Argentina^4^.

Patients with COVID-19 present an eminently respiratory affectation that can trigger in its most severe forms an acute respiratory distress syndrome, although cardiac, hematological, digestive, neurological and renal affectation has also been described^5–7^. The incidence of acute kidney injury (AKI) in patients hospitalized for COVID-19 varies between 0.5% and 80.3% according to the different series^5–12^. Direct viral cytotoxicity in kidney cells and kidney damage secondary to hypoperfusion and hypoxia due to hemodynamic instability, heart and lung damage, damage mediated by pro-inflammatory cytokines such as thrombotic microangiopathy and rhabdomyolysis, among others, have been described as possible etiologies of AKI^13-15^. As in AKI due to other causes, AKI secondary to COVID-19 has been associated with adverse outcomes, including worsening of other comorbidities, greater use of health care resources, and higher mortality^16^. Current reports of mortality among subjects hospitalized for COVID-19 with AKI vary widely across different cohorts, ranging from 7% to 100%^12,17–19^. Factors associated with AKI and its impact on patients hospitalized for COVID-19 have not yet been evaluated in our country. Local studies are needed to better understand the AKI associated with COVID-19 and thus optimize the management of this disease and its complications.

The objectives of the study were to evaluate the incidence, risk factors, and impact of acute renal failure in hospitalized subjects due to active SARS-CoV-2 infection in two third level hospitals in Córdoba, Argentina.

## Material and methods

A retrospective cohort study was conducted. Adults who were hospitalized due to active SARS-CoV-2 infection between March 3, 2020 and October 31, 2020 in the Hospital Privado Universitario de Córdoba and Hospital Raúl Angel Ferreyra were consecutively included.

Patients hospitalized for reasons other than COVID-19 and those who were referred to another institution within the first 48 hours of admission were excluded.

An active SARS-CoV-2 infection was defined as: confirmed diagnosis by any of the available methods (rT-PCR, antigen or serology for SARS-CoV-2) and admission to hospital within the first 21 days from the onset of symptoms or confirmation of the diagnosis (whichever occurred first).

Demographic data, comorbidities, clinical characteristics, and complementary studies carried out at hospital admission, as well as those related to the treatment and evolution of each patient, were obtained from the electronical medical record. Each case was followed up throughout the hospitalization.

AKI was defined as an increase in serum creatinine by ≥0.3 mg/dL in 48 hours, or an increase in serum creatinine ≥1.5 times the baseline value, which is known or presumed to have occurred in the previous seven days, or urine volume <0.5 ml/kg/hour for six hours^20^. The stages of AKI were defined as: Stage 1-increase in serum creatinine 1.5 to 1.9 times the baseline value or increase in serum creatinine by ≥0.3 mg/dL or decrease in urinary volume <0.5 ml/kg/hour for 6 to 12 hours. Stage 2-increase in serum creatinine from 2.0 to 2.9 times the baseline value or decrease in urinary volume <0.5 ml/kg/hour for ≥12 hours. Stage 3-increase in serum creatinine to 3.0 times the baseline value or increase in serum creatinine to ≥4.0 mg/dL or decrease in urine volume to <0.3 ml/kg/hour for ≥24 hours or anuria during ≥12 hours or requirement of renal replacement therapy (RRT)^20^.

For statistical analysis, continuous variables were expressed as mean and standard deviation or median (M) and 25-75% percentile interquartile range (IQR) and their comparison was made with Student’s t-test or Mann-Whitney according to the variable distribution. Categorical variables were expressed as absolute frequency (n) and relative frequency (%) and were compared with chi-square test or Fisher’s exact test, according to the expected frequencies. For the multivariate analysis, multiple logistic regression was used and all the variables that had been statistically significant in the univariate analysis were included in the model.

Adjusted Odd ratio (aOR) with its 95% confidence interval (95% CI) was used as association measures. All the tests were two-tailed and a *p* value less than 0.05 was considered statistically significant. Statistical analysis was performed with Stata 14 statistical program (Stata-Corp. LP., College Station, TX, USA).

The study was reviewed and approved by the research committee of the Hospital Privado Universitario de Córdoba.

## Results

During the study period, 450 adults were hospitalized due to active SARS-CoV-2 infection. Two patients were referred to another institution during the first 48 hours and were excluded from the analysis (Figure 1). A total of 448 patients were included, with a median age of 63 years (IQR 53-75). 63.6% were men. 17% had criteria for admission to the intensive care unit (ICU) at hospital admission. The most frequent comorbidities were hypertension (55.1%), obesity (31.7%) and diabetes (28.1%) and the most frequent clinical manifestations were fever (74.8%) and respiratory symptoms (cough 66.5% and dyspnea 60.7%). Pneumonia was evidenced in 95.8% of the patients (Table 1). The incidence of AKI was 19% (n=85/448). Of these, 36 (42.3%) presented AKI upon admission and 49 (57.7%) developed it during hospitalization. 50.6% presented AKI stage 1 (n 43), 20% stage 2 (n 17) and 29.4% stage 3 (n 25). In this last group, 18 subjects required RRT (figure 1). The incidence of AKI among patients who remained only in general ward throughout their hospitalization was 10.8% (n=31/286) and among subjects who required ICU at some point during their hospitalization was 33.3% (n=54/162).

**TABLE 1.** Baseline characteristics and clinical presentation at the time of hospital admission of the studied population.

| Baseline clinical characteristics | Todos (n 448) |
| --- | --- |
| Institution |  |
| Hospital Privado Universitario de Córdoba, n (%) | 239 (53.3) |
| Hospital Raúl Angel Ferreyra, n (%) | 209 (46.7) |
| Meets admission criteria in |  |
| General ward, n (%) | 372 (83) |
| Intensive care unit, n (%) | 76 (17) |
| Demographic characteristics |  |
| Male gender, n (%) | 285 (63.6) |
| Age, M (IQR) | 63 (53-75) |
| Comorbidities |  |
| Arterial hypertension, n (%) | 247 (55.1) |
| Obesity, n (%) | 142 (31.7) |
| Diabetes mellitus, n (%) | 126 (28.1) |
| Chronic kidney disease, n (%) | 46 (10.3) |
| Immunocompromised, n (%) | 43 (9.6) |
| Ischemic heart disease, n (%) | 35 (7.8) |
| Chronic heart failure, n (%) | 31 (6.9) |
| Clinical manifestations at hospital admission |  |
| Fever, n (%) | 335 (74.8) |
| Cough, n (%) | 298 (66.5) |
| Dyspnea, n (%) | 272 (60.7) |
| Asthenia, n (%) | 231 (51.6) |
| Myalgia, n (%) | 131 (29.2) |
| Diarrhea, n (%) | 106 (23.7) |
| Laboratory analysis at hospital admission |  |
| Leukocytes/μL, Median (IQR) | 7000 (5200-9675) |
| Neutrophils/μL, Median (IQR) | 5176 (3593-7480) |
| Lymphocytes/μL, Median (IQR) | 1029 (710-1452) |
| Platelets less than 110,000/mm<sup>3</sup>, n (%) | 37(8.3) |
| Creatinine in mg/dL, Median (IQR) | 0.99 (0.79-1.22) |
| Urea in mg/dL, Median (IQR) | 35 (27-48) |
| C-reactive protein in mg/dL, Median (IQR) | 7.01 (3.67-12.1) |
| Procalcitonin in ng/mL, Median (IQR) (performed in only 204 patients) | 0.13 (0.08-0.25) |
| Ultrasensitive troponin T in ng/L, Median (IQR) (253 studies were conducted) | 10.4 (6.45-21.9) |
| Ferritin in ng/dL, Median (IQR) (Only performed in 431 patients) | 1033 (510-1893) |
| D-dimer in μg/mL, Median (IQR) (only performed in 418 patients) | 0.55 (0.27-1.11) |
| Complementary images |  |
| With pneumonia, n (%) | 429 (95.8) |

*IQR= interquartile range*

**Figure 1.**
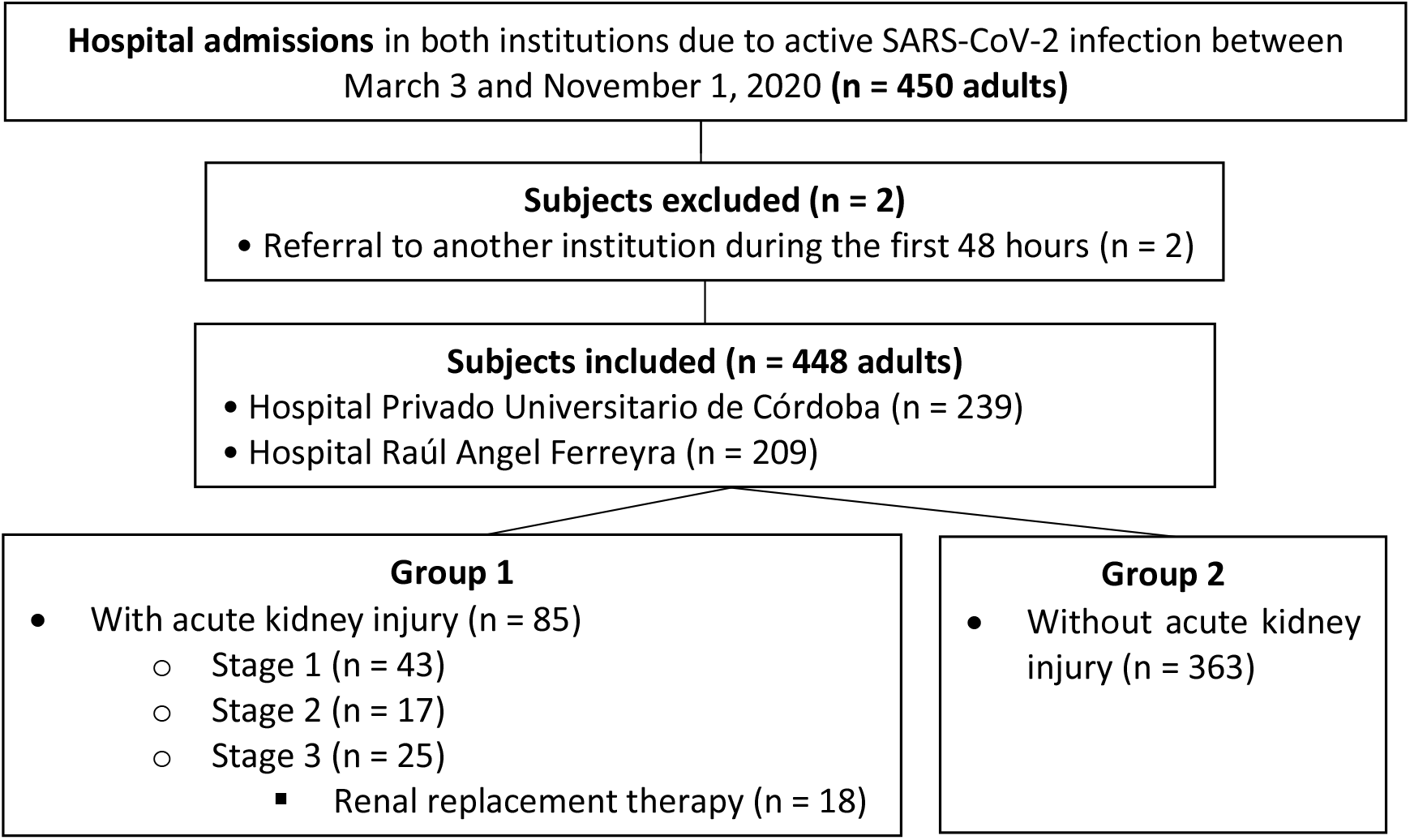
Flow chart of the inclusion of the subjects in the study and the assignment of groups.

Regarding patients with AKI, a higher percentage had criteria for admission in ICU, were predominantly males and were older, compared to those who did not present AKI (control group). As for comorbidities, the AKI group had a higher percentage of chronic kidney disease (CKD), immunocompromise, and ischemic heart disease compared to the control group. In the admission laboratory tests, subjects with AKI had higher values of total leukocytes, neutrophils, higher percentage of thrombocytopenia, higher levels of procalcitonin, troponin T and dimer D. All these characteristics were considered risk factors and variables significantly associated with AKI in the univariate analysis. There was also a clear trend for patients with AKI to have a higher percentage of arterial hypertension and chronic heart failure as comorbidities, lymphopenia at admission, and higher levels of C-reactive protein and ferritin, but these differences were not statistically significant (Table 2).

**TABLE 2.** Differences between baseline characteristics and clinical presentation at hospital admission between patients with and without acute kidney injury (AKI).

| Baseline clinical characteristics | AKI + (n 85) | AKI - (n 363) | p |
| --- | --- | --- | --- |
| Institution |  |  |  |
| Hospital Privado, n (%) | 49 (57.6) | 190 (52.3) | 0.377 |
| Hospital Raúl Angel Ferreyra, n (%) | 36 (42.3) | 173 (47.7) |  |
| Meets admission criteria in |  |  |  |
| General ward, n (%) | 58 (68.2) | 314 (86.5) | <0.001 |
| Intensive care unit, n (%) | 27 (31.8) | 49 (13.5) |  |
| Demographic characteristics |  |  |  |
| Male gender, n (%) | 62 (72.9) | 223 (61.4) | 0.047 |
| Age, M (IQR) | 69 (62-77) | 62 (52-74) | 0.001 |
| Comorbidities |  |  |  |
| Arterial hypertension, n (%) | 53 (62.3) | 194 (53.4) | 0.137 |
| Obesity, n (%) | 27 (31.8) | 115 (31.7) | 0.988 |
| Diabetes mellitus, n (%) | 26 (30.6) | 100 (27.6) | 0.575 |
| Chronic kidney disease, n (%) | 25 (29.4) | 21 (5.8) | <0.001 |
| Immunocompromised, n (%) | 13 (15.3) | 30 (8.3) | 0.048 |
| Ischemic heart disease, n (%) | 14 (16.5) | 21 (5.8) | 0.001 |
| Chronic heart failure, n (%) | 9 (10.6) | 22 (6.1) | 0.139 |
| Clinical manifestations at hospital admission |  |  |  |
| Fever, n (%) | 62 (72.9) | 273 (75.2) | 0.665 |
| Cough, n (%) | 60 (70.6) | 238 (65.6) | 0.377 |
| Dyspnea, n (%) | 51 (60) | 221 (60.9) | 0.881 |
| Asthenia, n (%) | 42 (49.4) | 189 (52.1) | 0.659 |
| Myalgia, n (%) | 27 (31.8) | 104 (28.6) | 0.570 |
| Diarrhea, n (%) | 21 (24.7) | 85 (23.4) | 0.801 |
| Laboratory analysis at hospital admission |  |  |  |
| Leukocytes/μL, M (IQR) | 8400 (5500-10800) | 6900 (5100-9000) | 0.009 |
| Neutrophils/μL, M (IQR) | 6003 (3900-8827) | 5103 (3534-7227) | 0.028 |
| Lymphocytes/μL, M (IQR) | 882 (621-1407) | 1075 (736-1464) | 0.064 |
| Platelets less than 110,000/mm<sup>3</sup>, n (%) | 16 (18.8) | 21 (5.8) | <0.001 |
| Creatinine in mg/dL, M (IQR) | 1.35 (1.06-1.83) | 0.93 (0.74-1.11) | <0.001 |
| Urea in mg/dL, M (IQR) | 53 (40-78) | 33 (25-44) | <0.001 |
| C-reactive protein in mg/dL, M (IQR) | 7.97 (4.1-13.85) | 6.68 (3.54-11.6) | 0.109 |
| Procalcitonin in ng/mL, M (IQR) | 0.22 (0.12-0.48) | 0.13 (0.07-0.2) | <0.001 |
| U-TT in ng/L, M (IQR) | 19.8 (10.4-37.2) | 8.9 (5.9-19.3) | <0.001 |
| Ferritin in ng/dL, M (IQR) | 1280 (652-1922) | 995 (470-1882) | 0.159 |
| D-dimer in μg/mL, M (IQR) | 0.7 (0.45-1.36) | 0.52 (0.24-1) | <0.001 |
| Complementary images |  |  |  |
| With pneumonia, n (%) | 80 (94.1) | 349 (96.1) | 0.378 |

AKI= acute kidney injury; M= Median; IQR= interquartile range; LDH= Lactate dehydrogenase; U-TT= Ultrasensitive troponin T

During hospitalization, 96.9% received prophylactic heparin, 85% dexamethasone or hydrocortisone, 57.6% antibiotics, and 37.5% convalescent plasma. A smaller percentage of patients received: hydroxychloroquine (0.7%), ritonavir/lopinavir (0.9%), nebulized ibuprofen (2.2%), and immunomodulatory drugs (1.1%). A higher percentage of subjects in the AKI group received antibiotics and convalescent plasma compared to the control group. The median length of hospital stay was 17 days (IQR=13.2-22). During follow-up, complications were: bacterial superinfection (12.3%), septic shock (8.8%), acute respiratory distress syndrome (ARDS, 27.9%), ICU admission (36.2%), mechanical ventilation (MV) requirement (14.7%). Overall mortality was 19%. Patients with AKI presented a longer hospitalization, greater requirement and time of stay in the ICU, a significant positive association with bacterial superinfection, sepsis, ARDS, MV and also presented higher mortality compared to the control group (mortality with AKI= 47.1% vs without AKI= 12.4%, table 3). Mortality incidence varied according to the stage of AKI (stage 1= 25.6%, stage 2= 41.2%, stage 3= 88%). In subjects who required RRT, all were on MV and mortality was 88.9% (n=16/18). The 2 survivors in this subgroup recovered their kidney function at the time of hospital discharge. In the adjusted multivariate analysis, the variables that were independently associated with AKI were: age (for every 10 years, aOR= 1.30, 95%CI=1.04-1.63, p=0.022), history of CKD (aORa=9.92, 95%CI=4.52-21.77, p<0.001), blood neutrophil count (per 1000 neutrophils, aOR=1.09, 95%CI=1.01-1.18, p=0.037) and MV requirement (aOR=6.69, 95%CI=2.24-19.9, p=0.001). And within the complications during follow-up, both AKI (aOR=3.32, 95%CI=1.6-6.9, p=0.001), ARDS (aOR=8.81, 95%CI=3.8-20.5, p<0.001) and MV (aOR=3.55, 95%CI=1.2-10.4, p=0.020) were independently associated with higher mortality.

**TABLE 3.** Medications received and associated complications in subjects with and without acute kidney injury (AKI).

| Events during follow-up | All (n 448) | AKI + (n 85) | AKI - (n 363) | p |
| --- | --- | --- | --- | --- |
| Medications received |  |  |  |  |
| Convalescent plasma, n (%) | 168 (37.5) | 43 (50.6) | 125 (34.4) | 0.006 |
| Corticosteroids, n (%) | 381 (85) | 71 (83.5) | 310 (85.4) | 0.663 |
| Antibiotics, n (%) | 258 (57.6) | 59 (67.4) | 199 (54.8) | 0.014 |
| Low molecular weight heparin /<br>Unfractionated heparin, n (%) | 434 (96.9) | 82 (96.5) | 352 (97) | 0.735 |
| Complications and events |  |  |  |  |
| Days of hospitalization, M (IQR) | 17 (13.2-22) | 21 (16-26) | 16 (13-22) | <0.001 |
| ICU requeriment, n (%) | 162(36.2) | 54 (63.5) | 108 (29.7) | <0.001 |
| Days in ICU, Median: IQR | 8 (4-15) | 12 (8-19) | 6 (3-11) | <0.001 |
| Mechanical ventilation, n (%) | 66 (14.7) | 42 (49.4) | 24 (6.6) | <0.001 |
| ARDS, n (%) | 125 (27.9) | 54 (63.5) | 71 (19.6) | <0.001 |
| Septic shock, n (%) | 39 (8.8) | 23 (27.4) | 16 (4.4) | <0.001 |
| Patients with bacterial<br>Superinfection, n (%) | 55 (12.3) | 32 (37.6) | 33 (9.1) | <0.001 |
| Death, n (%) | 85 (19) | 40 (47.1) | 45 (12.4) | <0.001 |

*AKI= acute kidney injury; M= Median; IQR= interquartile range; ICU= Intensive care unit; ARDS= Acute respiratory distress syndrome*

## Discussion

In the present study, the incidence, risk factors, and complications associated with AKI were evaluated in subjects hospitalized in two high complexity third-level hospitals in Córdoba, Argentina.

In a meta-analysis that included more than 13000 patients in China, the United States and different European countries, most of them hospitalized, the incidence of AKI was 17% and approximately 5% required RRT^21^. These percentages were very similar to those found in our study (AKI 19%, RRT 4%) and the criteria to define AKI were exactly the same. In Argentina, a single study carried out in a public tertiary hospital in Buenos Aires used the same AKI criteria^22^. In that prospective cohort they included 417 COVID-19 patients admitted to the general ward. AKI occurred in 4.8% of the patients, and 0.7% required RRT (only 3 patients, another 6 already required RRT prior to hospitalization). These percentages are much lower than those reported in our cohort and this may be mainly due to the fact that the subjects included were younger (median 43 vs 63 years of age), had fewer comorbidities such as hypertension (16.7% vs 55.1%), diabetes (15.3 % vs 28.1%) and CKD (3.1% vs 10.3%), the majority presented with a mild disease (86.8%) and a lower percentage of pneumonia (51.9% vs 95.8%), Patients with critical illness at admission were excluded in this study. Another series in Argentina reports an incidence of kidney injury of 7% but does not specify the criteria used to define AKI and both the baseline characteristics and the severity of the disease differ from that of the patients included in our cohort^23^.

Currently described independent predictors of AKI in subjects with COVID-19 include age, male gender, history of obesity, diabetes, hypertension, cardiovascular disease, CKD, elevated IL-6 levels, baseline ACEI-ARA-II use, exposure vancomycin and NSAIDs, septic shock, hypoxia, and MV^8-10,24,25^. In Argentina, to our knowledge there are no publications to date on the risk factors for AKI in patients with COVID-19. In our study, in addition to age, history of CKD and MV, blood neutrophil count at admission was also identified as an independent predictor of AKI, a variable that had not been identified in previous studies resulting from great value to consider in the initial evaluation. Furthermore, in the univariate analysis male sex, immunocompromise, coronary heart disease, thrombocytopenia, and higher inflammatory markers such as procalcitonin, troponin, and dimer-D were also associated with AKI, although later in the multivariate adjustment lost statistical significance. And other comorbidities such as hypertension and heart failure, a lower count of blood lymphocytes, and other inflammatory markers such as C-reactive protein and ferritin, presented a clear trend of association with AKI, but without statistical significance. This may be due to the number of subjects included and statistical power.

On the other hand, patients with AKI were more frequently treated with convalescent plasma and antibiotics; these differences are probably explained by the more severe condition that this group of patients presented and the higher percentage of bacterial superinfection.

In the same meta-analysis mentioned previously, the mortality rate of patients with AKI was 52%^21^, a percentage similar to that found in our study (47.1%). In our cohort, as in others, AKI has been identified as an independent predictor of mortality^9,24,26,27^, and the higher the AKI stage, the greater the risk^9^. In addition, AKI in our series was associated with other complications such as long-term hospitalization, admission to UCU and prolonged stay in ICU, among others.

The limitations of our study are mainly based on its retrospective nature, although in both institutions there was a protocol for action against COVID-19 which would tend to unify management.

This study is the first in our country to evaluate the incidence, risk factors, and prognosis associated with AKI in adults hospitalized for COVID-19. The epidemiological analysis of AKI incidence, degrees of affectation, requirement of ICU, MV and RRT can be useful for planning and allocating resources in the event of a possible new outbreak. The determination of AKI risk factors allows the early identification of the susceptible subjects. The analysis of complications and the impact associated with AKI, such as mortality and prolongation of hospital stay, reflect the importance of early diagnosis and adequate treatment.

In conclusion, the incidence of AKI in hospitalized subjects due to active COVID-19 infection was 19%. The independent predictors of AKI were age, CKD history, neutrophil count, and MV. AKI was associated with bacterial superinfection, sepsis, respiratory distress syndrome, prolonged hospitalization days, higher ICU and MV requirement, and was also independently associated with higher in-hospital mortality.

## Data Availability

The data was deidentified by the investigator and is restricted to use by authorized individuals. The deidentified dataset is not publicly available.

## Acknowledgments

To the “Fundación Nefrológica de Córdoba” and “Instituto Universitario de Ciencias Biomédicas de Córdoba (IUCBC)” for the support to cover publication costs.

## Conflict of interest

None to declare.

## KEY POINTS

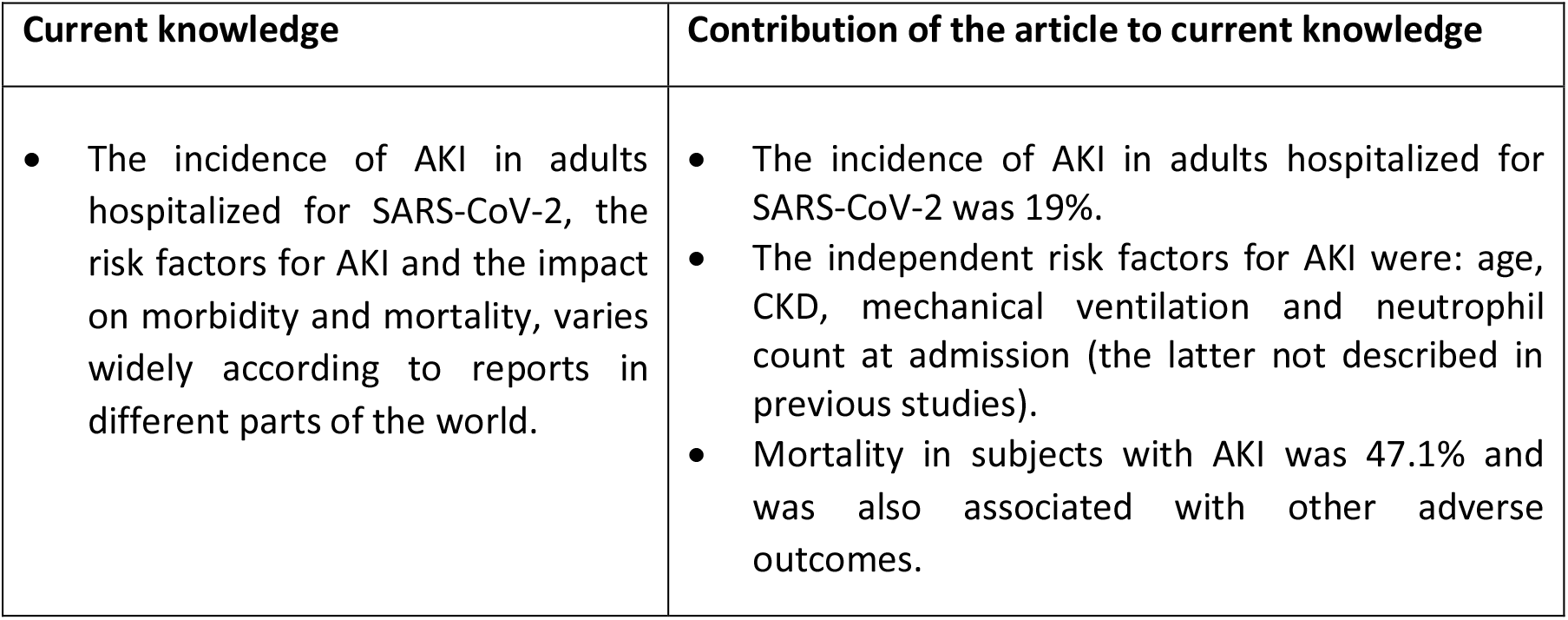

## Notes

### Competing Interest Statement

The authors have declared no competing interest.

### Funding Statement

No external funding was received

### Author Declarations

Ethical oversight was waived by the Institutional Committee of Ethics in Health Research (C.I.E.I.S) of the Hospital Privado Universitario de Cordoba (Cordoba, Argentina) given the observational characteristic of the study and the de-identified of the data. The study was reviewed and approved by the research committee of the Hospital Privado Universitario de Cordoba, Argentina.

